# Survey of Attitudes on Personal Protection Interventions Against COVID-19 Including MMR Vaccination and Future Anti-COVID Vaccines

**DOI:** 10.1101/2020.10.21.20215251

**Authors:** Joseph D. Schulman, James N. Cooper, Gary W. Crooks

## Abstract

An electronic survey was conducted in October, 2020 among individuals primarily age 60 and older regarding their degree of confidence of deriving personal protection from 8 different anti-COVID interventions – social isolation, lockdowns, avoiding restaurants, taking MMR vaccine, wearing masks when indoors with others, avoiding hotels, avoiding commercial air travel, and using the first future specific anti-COVID vaccine. Responses were received from 135 persons from many different U.S. regions and 5 foreign countries. Respondents were generally individuals with very high levels of education and personal achievement. Results demonstrated wide diversity of responses regarding each of these interventions. None were strongly supported by a majority of respondents, but those receiving the largest proportions of strong support were social isolation (41%), wearing masks indoors (41%), and using the first anti-COVID vaccine (41%). MMR (measles-mumps-rubella) vaccination was viewed much more positively than negatively but had the highest proportion of individuals who felt they had insufficient data to formulate an opinion. The largest number of strong negative assessments were toward lockdowns (37%). We speculate that the wide variation in perception of possible benefits from the surveyed interventions, most of which have been widely practiced by or imposed upon millions of individuals, in this highly accomplished older population at increased personal risk from COVID-19 reflects the current absence of rigorous scientific proof of the efficacy of any these interventions, and the continuation of the epidemic despite the widespread utilization of most of them.

## Introduction

The current COVID-19 epidemic due to SARS-CoV-2 is estimated to have caused over 1 million deaths worldwide^1^ and over 200,000 of these have been in the United States^1^. After approximately 8 months the epidemic continues to cause about 700 new deaths per day in the U.S.^1^ Public health authorities have made various recommendations on methods to reduce illness and death, and many states have imposed very strong restraints on people and businesses as part of this effort. We report here the results of a survey to assess the perceived likelihood of personal benefit from 6 commonly utilized interventions, the less widely recognized use of MMR vaccination, and a future initial specific anti-COVID vaccine.

## Methods

Approximately 400 persons were emailed a request to participate in the survey, and 135 people provided their responses in the week prior to October 15, 2020. The email distribution was intended to reach people who were almost all highly educated, high achievers, and primarily age 60 and above. The supplement indicates the wide variety of professional accomplishments of the survey responders. Individuals were asked to “indicate your degree of confidence that a particular anti-COVID intervention is likely to be useful for you personally in protecting yourself from illness or death due to COVID-19”. The degree of confidence level choices were the following: Less than 20%, 20-40%, 40-60%, 60 – 80%, greater than 80%, and unable to make an assessment. Respondents were also asked to state their age. The survey was accompanied by information on MMR vaccination^2,3,4^ and the proposed controlled clinical trial for MMR in 30,000 health care workers^5^. No information about other interventions was provided and all potential respondents were explicitly told that there was no “right” answer to any question except their age. All data was entered into and analyzed using Excel spreadsheets and the numerical results used to create graphs showing the distribution of answers regarding each intervention.

## Results and discussion

The results are shown in graphs 1-9. They show a remarkably varied degree of confidence of benefit for every intervention. Strikingly none of the interventions which have been widely utilized and in some cases forced upon their populations by state and city governments achieved even a majority of responses indicating a perceived high probability (80% or more) of personal benefit. This rating was given to social isolation by 41%, to lockdowns by 27%, to restaurant avoidance by 27%, to wearing masks indoors by 41%, to avoiding hotels by 26%, and to avoiding commercial air travel by 31%. Of the preceding 5 interventions the lowest perceived personal benefit rating went to lockdowns which 37% of respondents viewed as minimally beneficial to them if at all.

**Graph 1.**
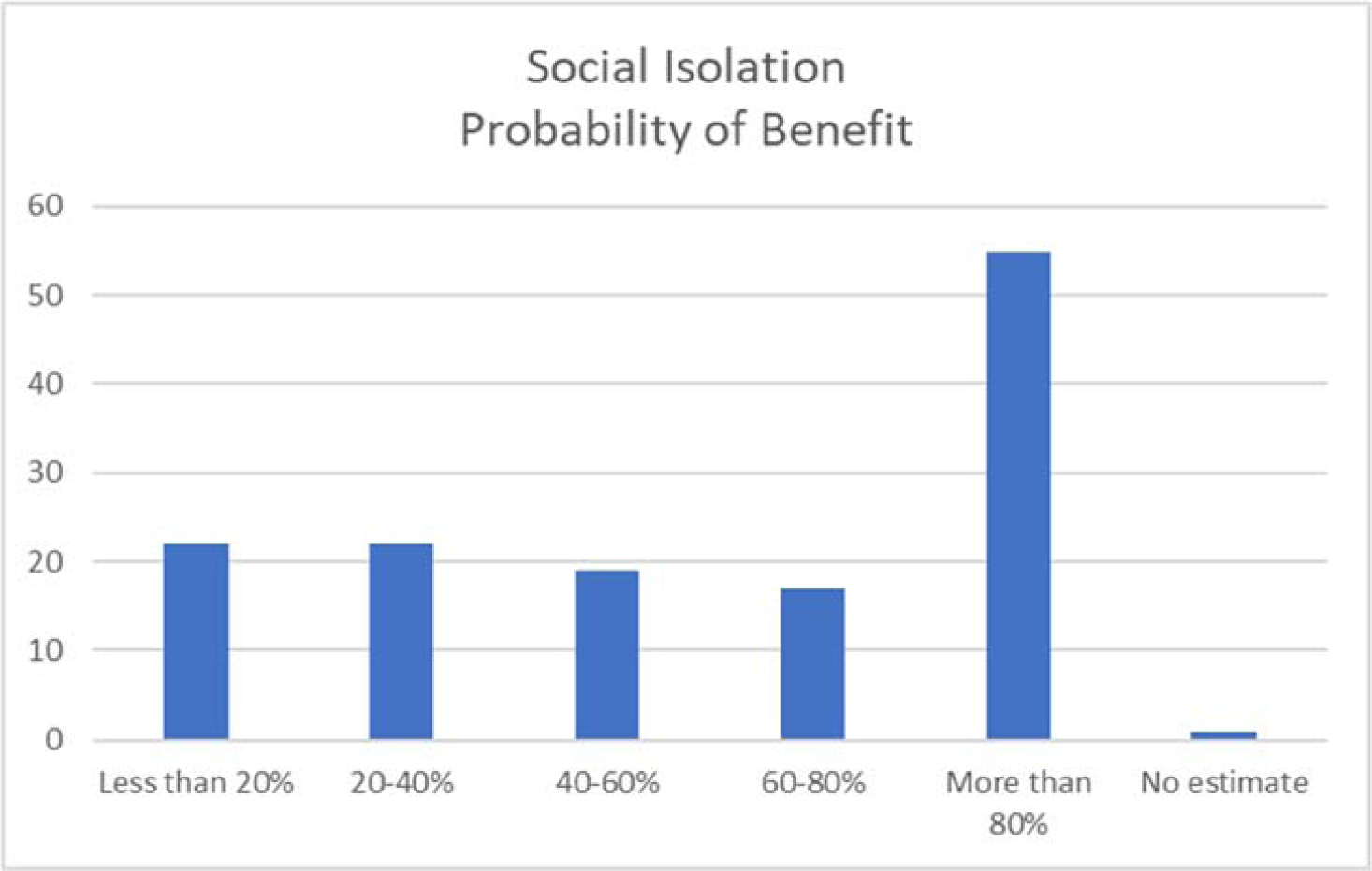
For all graphs the vertical axis is number of responses.

**Graph 2.**
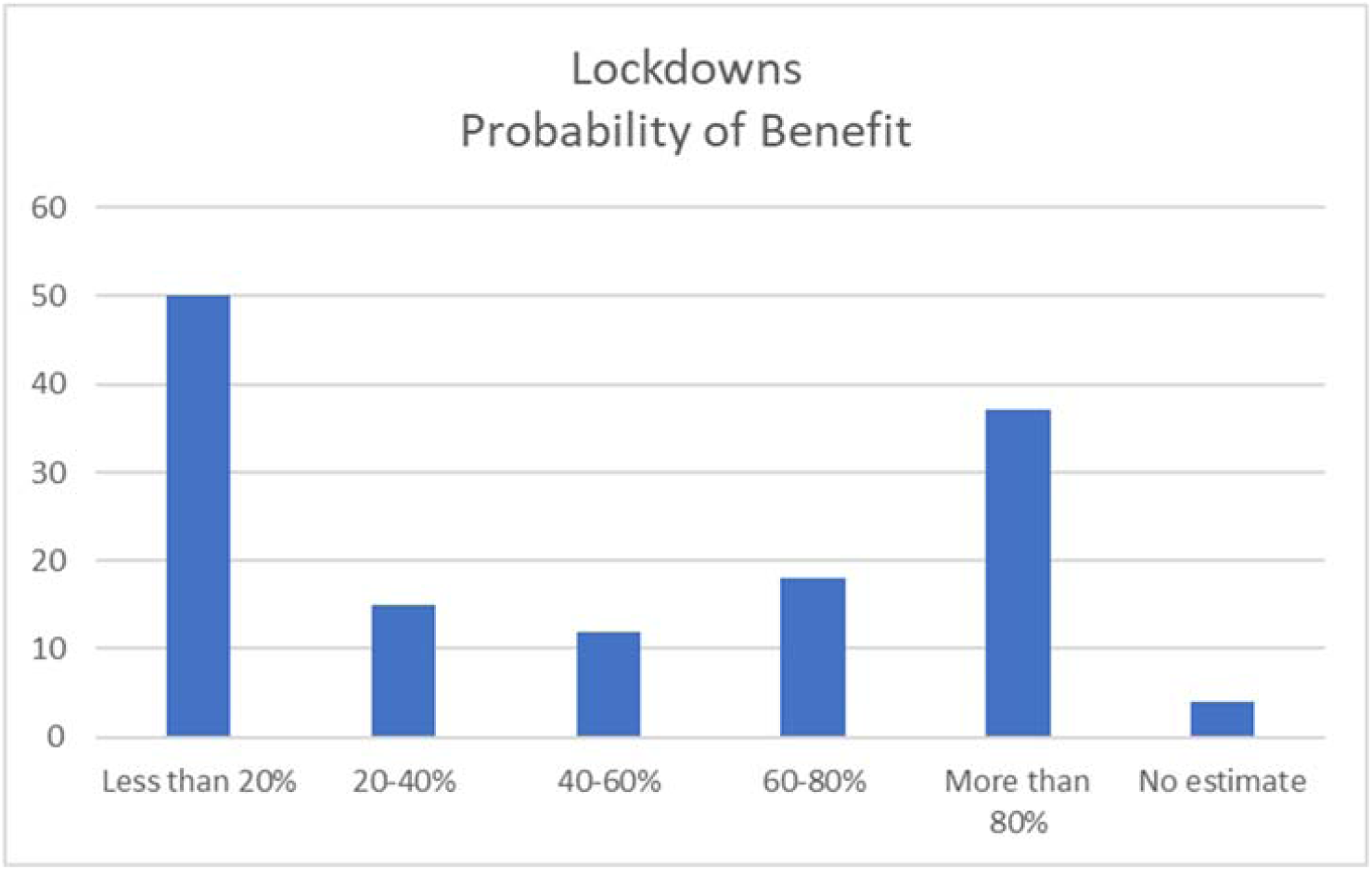

**Graph 3.**
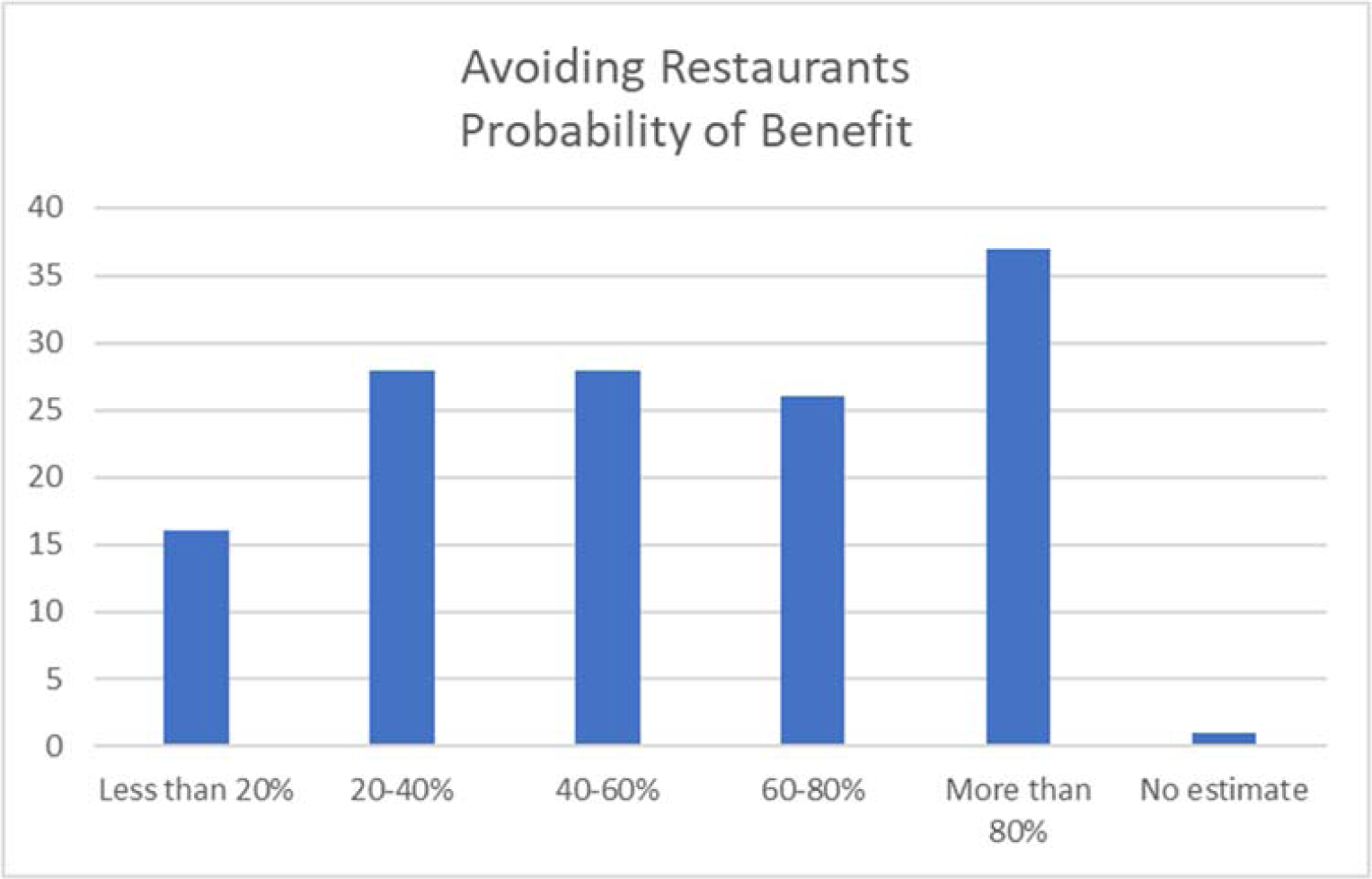

**Graph 4.**
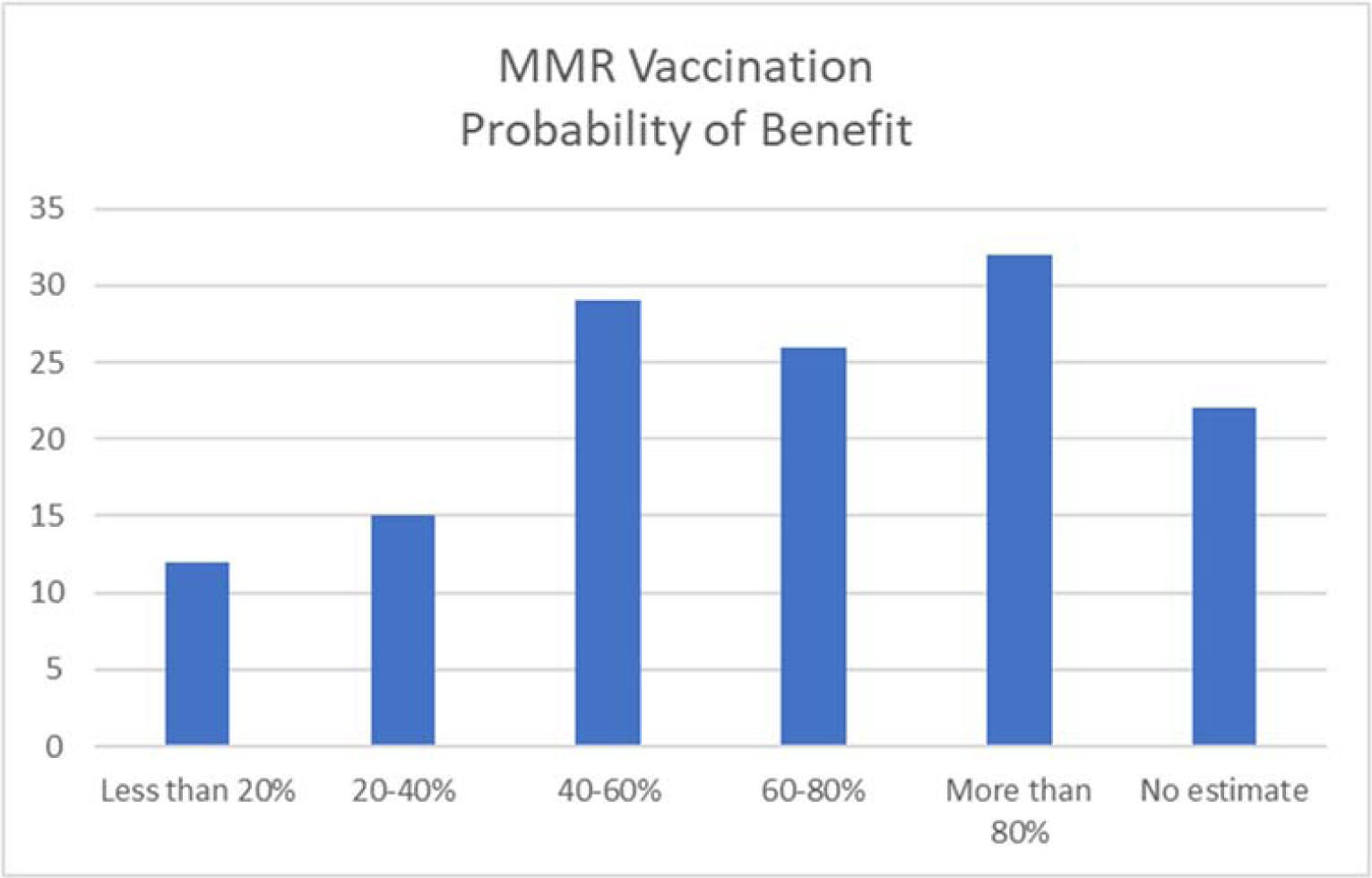

**Graph 5.**
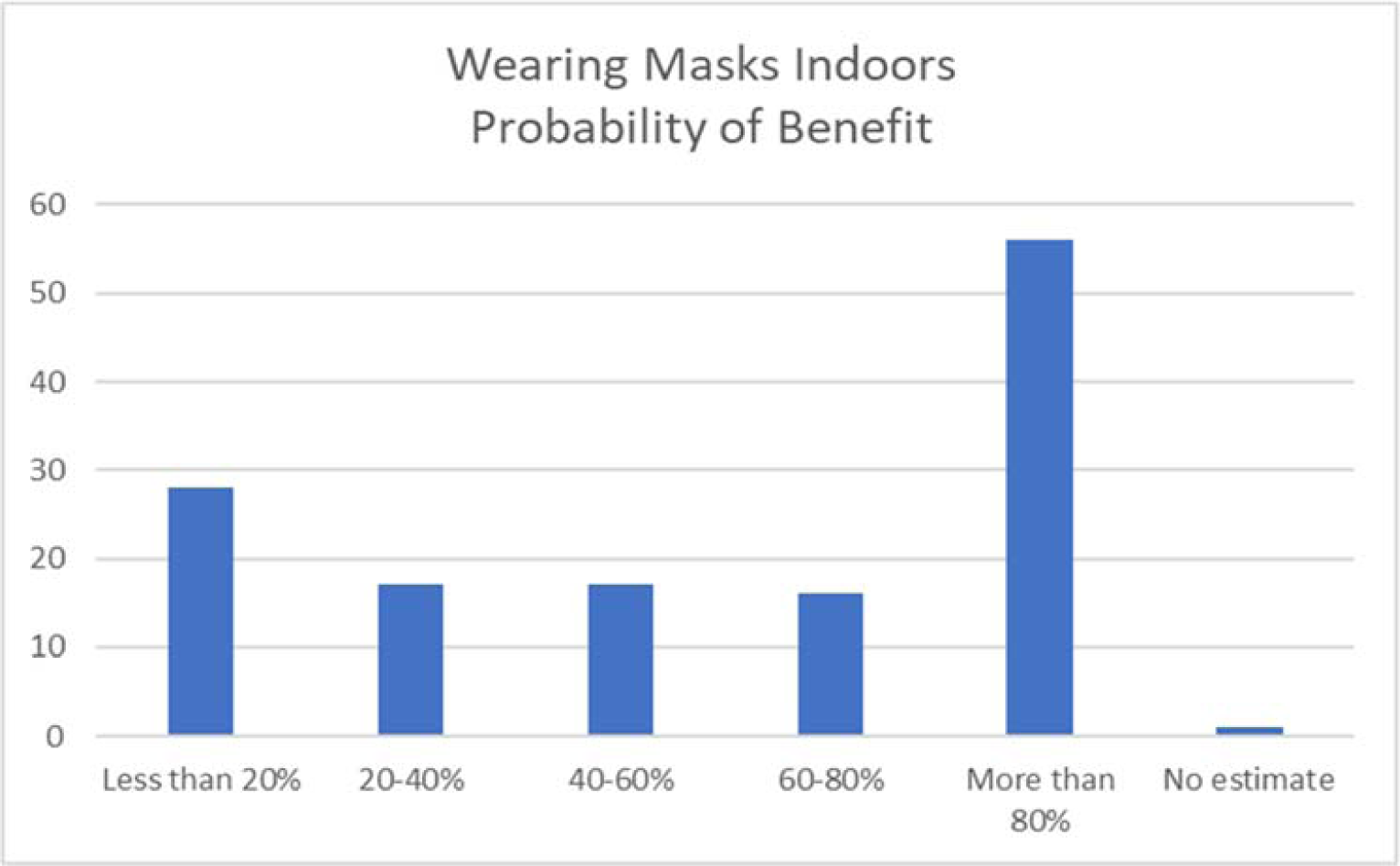

**Graph 6.**
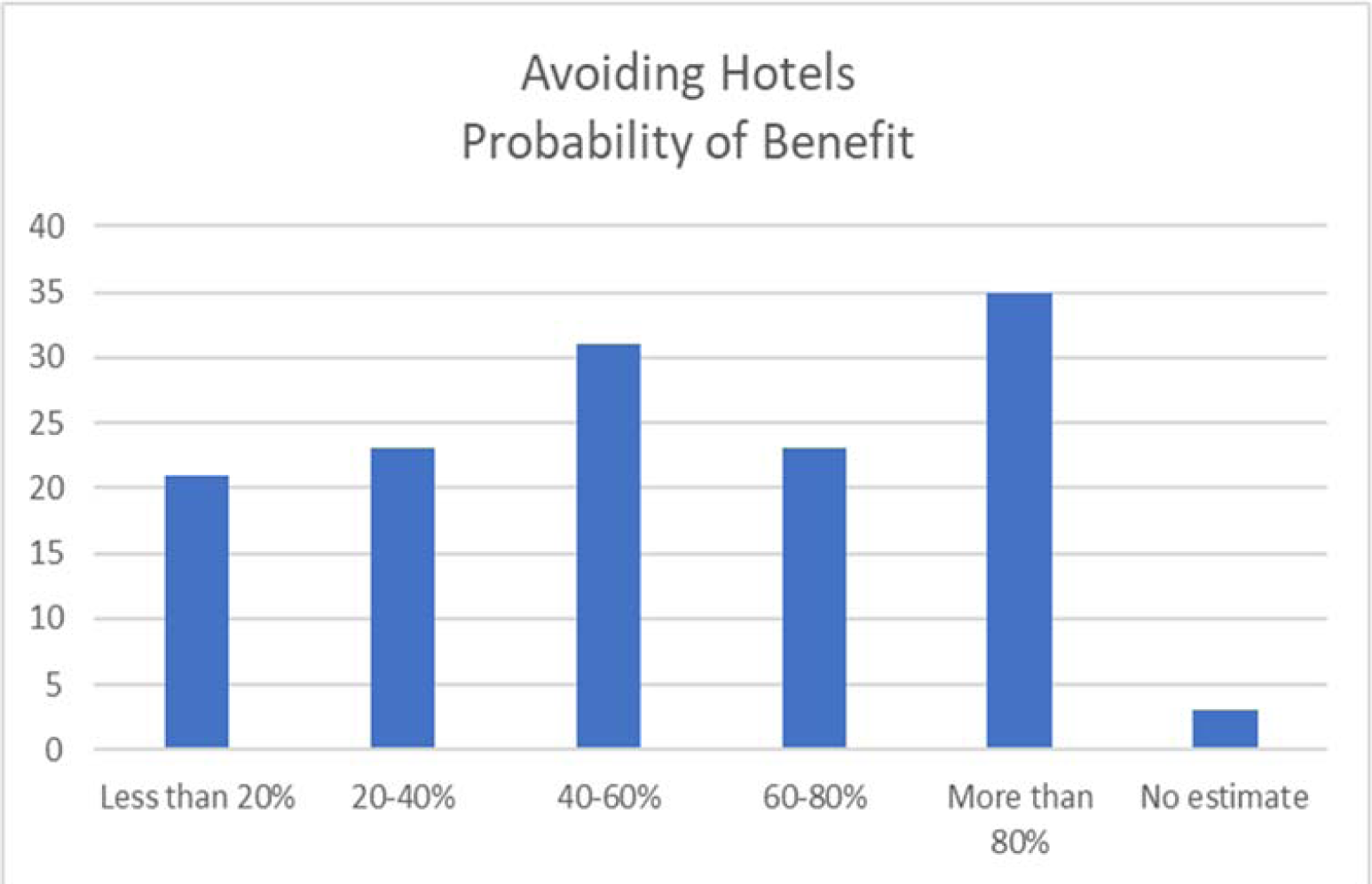

**Graph 7.**
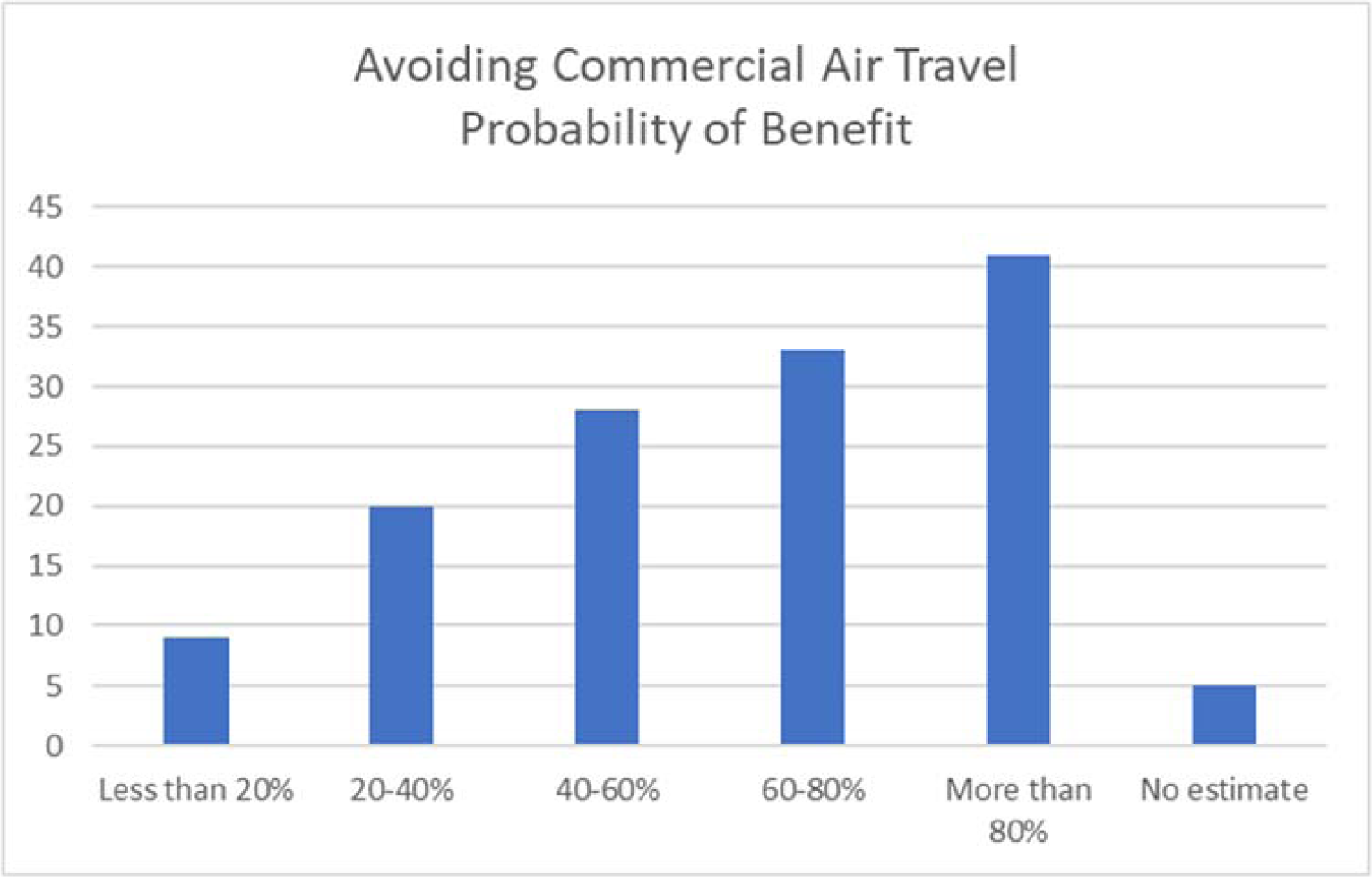

**Graph 8.**
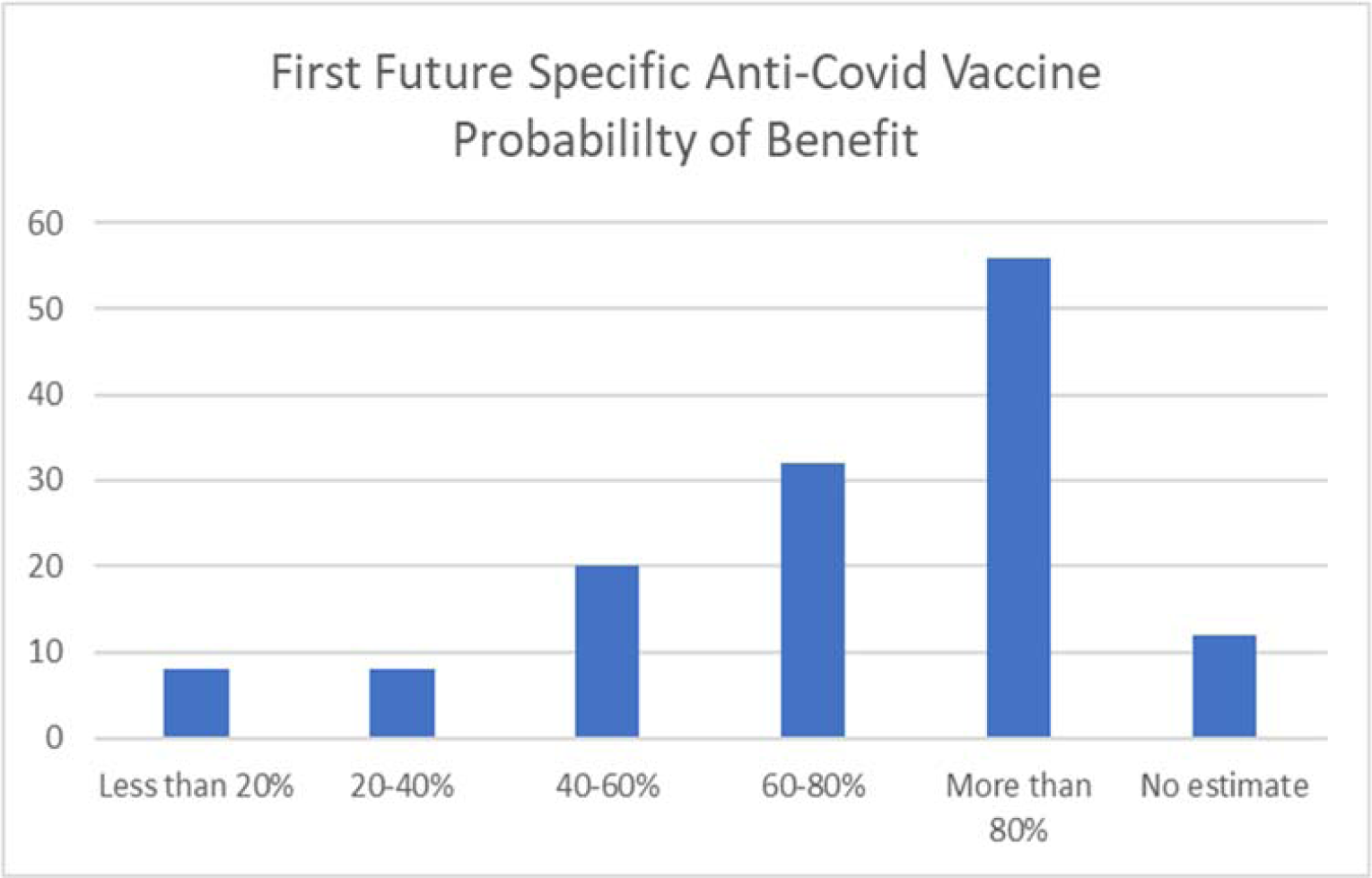

**Graph 9.**
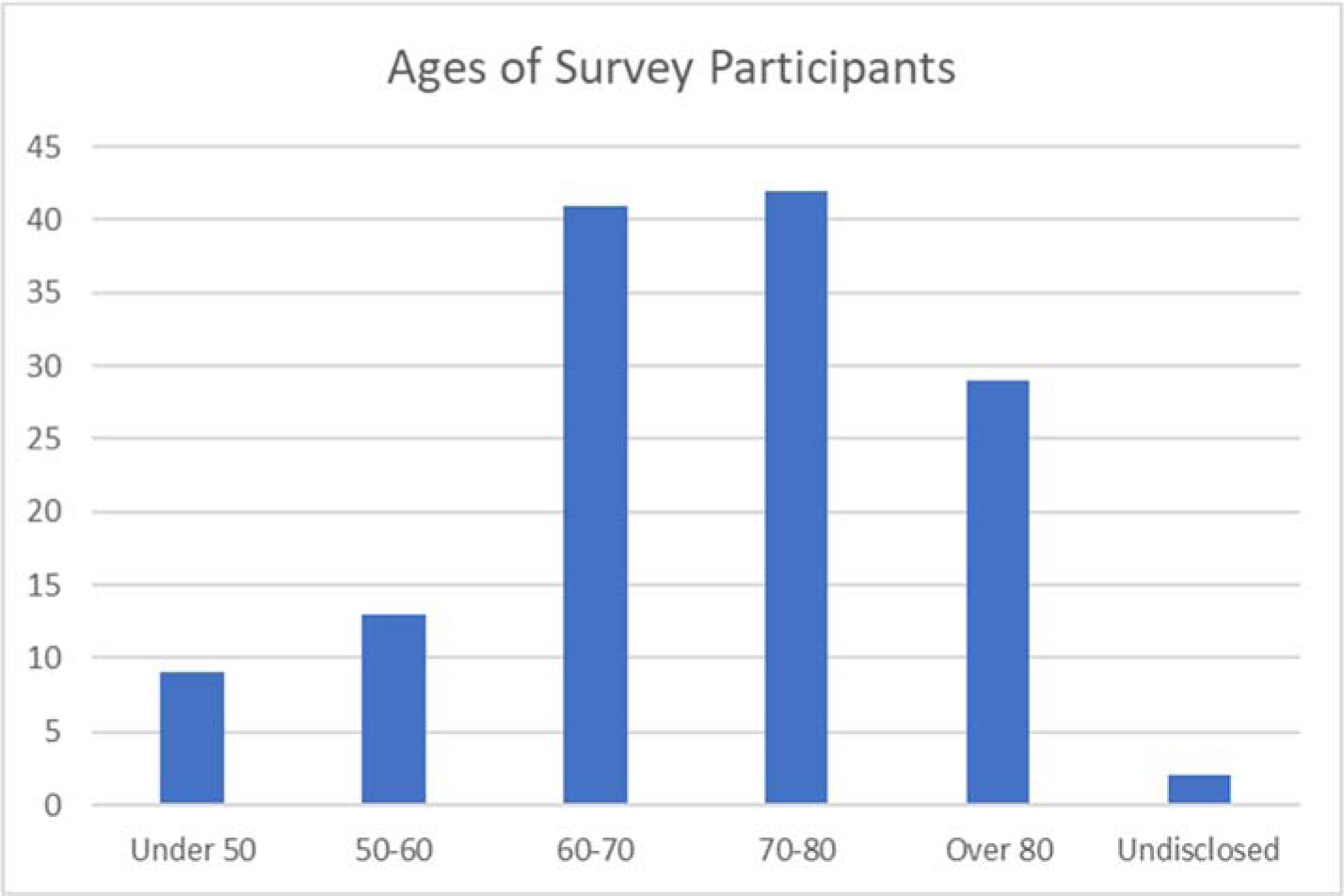

The results regarding MMR vaccination and use of a first future anti-COVID vaccine cannot be directly compared to the preceding 6 interventions which have been advocated to the public and in some cases been forced upon them for many months. MMR information was new to most of the survey recipients and was not comprehensive although it provided references to the most important work on this subject and described the proposed clinical trial. Not surprisingly a fairly high proportion of respondents (16%) felt unable to make an assessment of probability of personal benefit, a greater percentage than for any other intervention. The likely benefit of a future anti-COVID vaccine received the second largest number of answers of unable to make an assessment (9%). Nevertheless as shown in graphs 4 and 8 a far higher proportion of respondents were favorably inclined to currently using MMR vaccination or to future use of an anti-COVID vaccine than considered them highly unlikely to be of personal benefit.

Above are some highlights derived from the data, and the graphs are presented in full to permit others to draw their own conclusions. We will also provide full copies of the raw data linked to each numbered individual upon request to the corresponding author.

In conclusion we offer possible explanations for the results of this survey. In our opinion the remarkably wide dispersion of opinions on all surveyed interventions among a group of intelligent people whose age distribution (see graph 9) places them at much greater than average risk of illness and death from COVID-19 is substantially due to two major factors: (1) the lack of solid scientific proof of efficacy for any of them and (2) the experience of the last 8 months indicating that these interventions have failed to arrest the epidemic due to SARS-CoV-2. We believe very few members of the surveyed population would have ignored convincing proof of efficacy for any intervention if such proof existed. The implications of enforcing non-voluntary restrictions on millions of people absent such proof are beyond the scope of this inquiry.

## Data Availability

Copies of all data are available upon request from the corresponding author (JDS).

## Acknowledgments

We thank those who provided assistance with data reduction and made suggestions for improvement of this research.

## References

1. Worldometer website at https://www.worldometers.info/coronavirus/#countries

2. Gold, J.E., Tilley, L.P., Baumgartl, W.H. MMR vaccine appears to confer strong protection from COVID-19: few deaths from SARS-CoV-2 in highly vaccinated populations. Report published online via https://www.world.org/, May, 2020.

3. Young, A., Neumann, B., Mendez, R.F., et al. Homologous protein domains in SARS-CoV-2 and measles, mumps, and rubella viruses: preliminary evidence that MMR vaccine might provide protection against COVID-19. Preprint online at https://www.medrxiv.org/content/10.1101/2020.04.10.20053207v1 or https://doi.org/10.1101/2020.04.10.20053207

4. Sidiz, K., Dana, K., Shakhawan, A., and Kodius, R. Does early childhood vaccination protect against COVID-19? Front. Mol. Biosci., June 5, 2020 at https://www.frontiersin.org/articles/10.3389/fmolb.2020.00120/full

5. Washington University School of Medicine report at https://medicine.wustl.edu/news/global-trial-to-test-whether-mmr-vaccine-protects-front-line-health-care-workers-against-covid-19/

