## Supplemental Table 1 for "Survey of Attitudes on Personal Protection Interventions Against COVID-19 Including MMR Vaccination and Future Anti-COVID Vaccines"

**Supplement:**

At least 90% of respondents were from the United States and were widely dispersed having lived or worked in at least 30 different states including Maryland, Virginia, Florida, New York, Pennsylvania, California, Washington, Wyoming, and Colorado among many others. Responses were also received from Canada, Greece, Kuwait, Ireland, and Mexico. The respondents were, on average, a remarkably intelligent and accomplished group far above the population average, and highly diverse in their former or current professional fields. A partial list includes university presidents, professors in many disciplines both scientific and others such as business and history, state supreme court justices, founders or CEOs of public and private corporations, physicians in a variety of specialties, mathematicians, financial analysts, private investors, investment advisors, senior accountants, managing directors, insurance experts, engineers, architects, musicians, art experts, chief financial officers, senior administrators, heads of think-tanks, members of secret federal services, electricians, real estate professionals, software experts, club managers, public health experts, clinical laboratory directors, police chiefs, employment recruiters, airline pilots, patent experts, bankers, senior civil service employees, ministers, economists, dentists, security experts, golf professionals, farm owners, SEALs, movie producers, veterinarians, mayors, artists, corporate trainers, restaurateurs, media experts, charitable leaders, air traffic experts, materials experts, regulatory advisors, account executives, fundraisers, hospital administrators, contractors, and others.
